# HOW DO FIELD EPIDEMIOLOGISTS LEARN? A PROTOCOL FOR A QUALITATIVE INQUIRY INTO LEARNING IN FIELD EPIDEMIOLOGY TRAINING PROGRAMS

**DOI:** 10.1101/2023.12.05.23299419

**Authors:** Matthew Myers Griffith, Emma Field, Angela Song-en Huang, Tomoe Shimada, Munkhzul Battsend, Tambri Housen, Barbara Pamphilon, Martyn D. Kirk

**Author notes:** Corresponding author –.

## Abstract

**Introduction:** COVID-19 underscored the importance of field epidemiology training programs (FETP) as countries struggled with overwhelming demands. Experts are calling for more field epidemiologists with better training. Since 1951 FETP have been building public health capacities across the globe, yet explorations of learning in these programs are lacking. This qualitative study will 1) describe approaches to training field epidemiologists in FETP; 2) describe strategies for learning field epidemiology among FETP trainees; and 3) explain the principles and practices aligning training approaches with learning strategies in FETP.

**Methods and analysis:** The research design, implementation, and interpretation are collaborative efforts with FETP trainers. Data collection will include interviews with FETP trainers and trainees and participant observations of FETP training and learning events in four FETP in the Western Pacific Region. Data analysis will occur in three phases: I) we will use the constant comparison method of Charmaz’s grounded theory during open coding to identify and prioritise categories and properties in the data; II) during focused coding, we will use constant comparison and Polkinghorne’s analysis of narratives, comparing stories of prioritized categories, to fill out properties of those categories; III) we will use Polkinghorne’s narrative analysis to construct narratives that reflect domains of interest, identifying correspondence among Carr and Kemmis’s practices, understandings, and situations to explain principles and processes of learning in FETP.

**Ethics and dissemination:** We have obtained the required ethics approvals to conduct this research at The Australian National University (2021/771) and Taiwan’s Ministry of Health and Welfare (112206). Data will not be available publicly, but anonymised findings will be shared with FETP for collaborative interpretation. Ultimately, findings and interpretations will appear in peer reviewed journals and conferences.

**STRENGTHS AND LIMITATIONS OF THIS STUDY:**

- This study will be co-designed, co-developed, and co-interpreted with practitioners to generate relevant, useful, and informative findings for field epidemiology training programs, practitioners, and learners.
- Use of multiple data collection methods and theoretical frameworks will improve the credibility of the findings.
- Engagement of participants and programs throughout the process to check interpretations and facilitate dialogue on findings will strengthen the trustworthiness of the findings.
- The methodology aims to explore experiences in depth, and resources restrict the number of programs and participants that may enrol. So, there will be limits to the generalizability of the findings beyond the included programs.
- As grounded theory aims for hypothesis generation not hypothesis testing, the findings will be limited to explanations of training and learning and thus not interpretable as statements of the effectiveness of training approaches or programs.

## INTRODUCTION

COVID-19 highlighted the importance of field epidemiology training programs (FETP).(1-4) Nevertheless, many countries struggled to cope with the demands of the pandemic. These struggles have been linked to insufficient numbers of, inadequate training for, and low government regard for field epidemiologists.(5)

FETP “provide critically needed public health and global health security services through a mentored, learn-by-doing approach that emphasizes fieldwork and improves the effectiveness of the workforce and the systems required to provide those services”.(6) Training includes 25% classroom and 75% field experiences (Figure 1). Core topics include outbreak investigation, public health surveillance, epidemiologic methods, data management and analysis, and public health communication.(6, 7)

**Figure 1.**
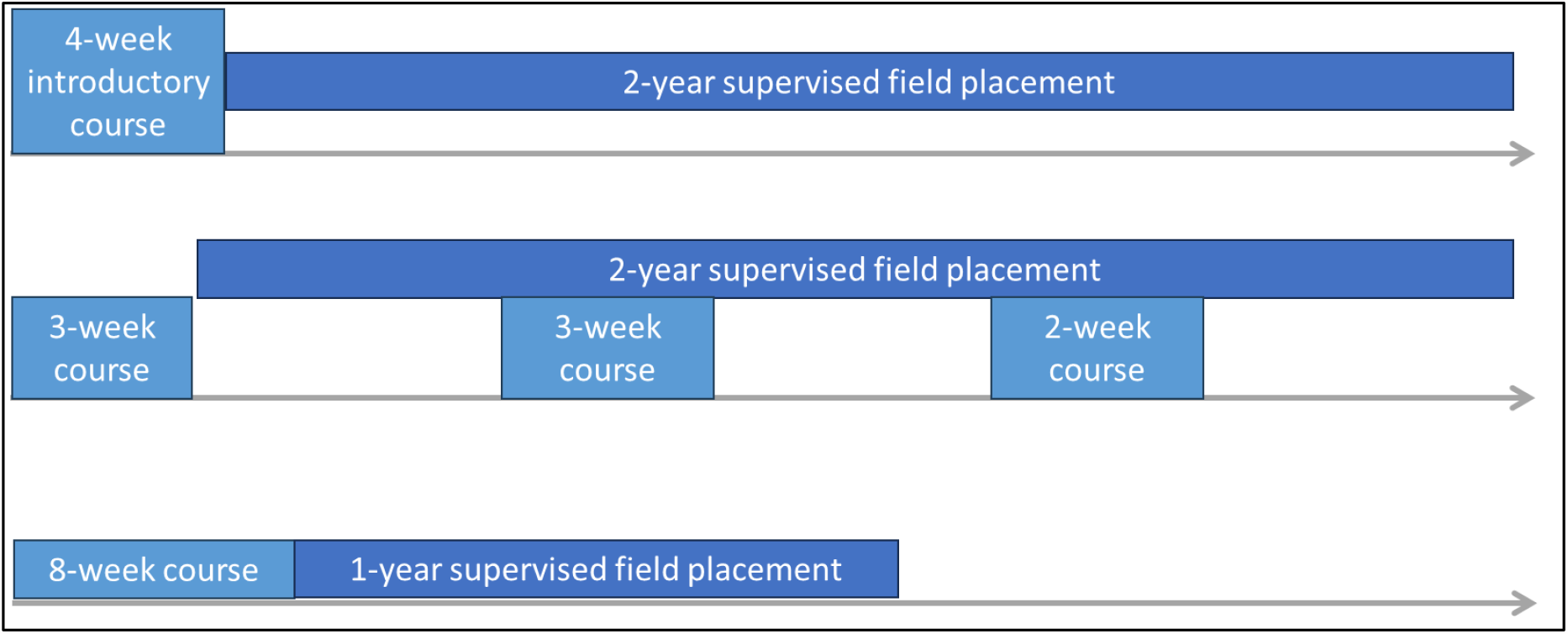
Example FETP designs. (Top) a 2-year “Advanced” program with intensive introductory course like the programs in japan and Taiwan; (middle) a 2-year “Advanced” program with module-based instruction like Australia’s program; (bottom) a 1-year “Intermediate” program with intensive introductory course like the program in Mongolia.

FETP have been building capacity for over 70 years. In 1951, Alexander D. Langmuir created the Epidemic Intelligence Service (EIS), aiming to develop epidemic expertise by turning medical clinicians into field-savvy epidemiologists, transforming their focus from the individual patient to the population.(8, 9) The first EIS class had twenty-two White male physicians and one sanitary engineer.(10) In the 1970s and 1980s, countries began to copy the EIS model.(11, 12) Today, 98 programs build public health workforce capacities in more than 200 countries and territories.(13)

Despite FETP’s importance, longevity, and spread, explanations of its learning processes are lacking. One review of the FETP literature notes an impressive list of outputs with few discussing how trainees apply new knowledge, skills, or attitudes.(14) Recent work continues to document FETP outputs and contributions to public health.(1, 3, 4) Although some publications utilize Kirkpatrick’s (15) training evaluation levels to assess application of learning, they do not explore the learning process.(16, 17) For instance, an evaluation of multiple FETP focused on Kirkpatrick’s levels 3 (behaviour) and 4 (results) found that most graduates engaged in field epidemiology activities and perceived their FETP experience to have helped them perform these roles. It did not assess program competencies or skills of the graduates nor explicate the learning process.(18)

In contrast, articles describing learning processes in medicine and nursing abound.(19-24) To take one example, Ohta and colleagues utilized grounded theory to reveal learning processes in Japanese medical students’ transitions to rural community hospitals.(25) They found that integration of cognitive apprenticeship (26) and legitimate peripheral participation in communities of practice (27) with learners’ regular reflection on performance facilitated their learning family medicine. For a second example in an outbreak-like setting, Taber and colleagues illuminate learning processes of paramedics and firefighters in Canada.(28) Their findings align with Lave and Wenger’s (27) learning characterizations. Training sessions are only a beginning, while understanding comes in practice where they learn from one another and adapt to fit their realities into the policies, protocols, and structures to respond to emergencies. When they face novel situations, paramedics and firefighters employ active, creative, and immediate learning that the authors note merit further research. No such studies appear for learning in ‘field epidemiology’ or ‘epidemiology’.

Considering the importance, diverse contexts in which it operates, and perceived success at training public health professionals, FETP is fertile ground for learning research. We propose a qualitative exploration of learning in FETP with the following objectives:

1. Describe approaches to training field epidemiologists in field epidemiology training programs.
2. Describe strategies for learning field epidemiology among trainees in field epidemiology training programs.
3. Explain the principles and practices that align training approaches with strategies for learning in field epidemiology training programs.

We anticipate that understanding the FETP learning principles and practices will inform how to upscale, adapt to context, balance standardization and adaptation, and measure learning and how training approaches should evolve with the theories informing adult learning today. Also, we hope that this protocol informs future research into FETP learning across diverse geographical, wealth, and program contexts.

## METHODS AND ANALYSIS

### Study design

This qualitative, exploratory research will be designed, implemented, and interpreted with end-users of the findings. The germ for research into FETP learning came with the principal investigator (PI), but the design, including the questions and analytical approach, and the interpretation of data will be collaborative efforts with FETP trainers of participating programs who meet virtually each month, following the Critical Reference Group approach of Wadsworth. Involving trainers in this collaborative approach is expected to improve the study’s relevance for the people who share the problem; focus research questions; enhance study relevance for those whose jobs entail doing something about it; increase the research design’s effectiveness; improve the meaningfulness of the information gathered; strengthen power and accuracy of the theory that the research generates; improve relevance, creativity, and effectiveness of the actions that are based on the study; and strengthen commitments for following up on actions and researching them further.(29)

We will use two methods for collecting data: participant observations of FETP training and learning events and in-depth unstructured interviews with FETP trainers and trainees. We will analyse the data in three phases, employing the constant comparison method of grounded theory (30, 31) during the first two phases to generate categories and properties inductively from the data. We will employ analysis of narratives and narrative analysis (32) during the second two phases to construct narratives that reflect the domains of interest. In the third phase, we will integrate the narratives to identify correspondence and non-correspondence among practices, understandings, and situations (33) to explain the principles and processes of learning in FETP.

### Reflexivity Statement

The PI and co-researchers of this study are public health professionals who have worked in governments, clinics, hospitals, universities, WHO, US CDC, and FETP. We were born, raised, and educated in multiple countries, mostly in the Western Pacific Region. Most of us are alumni of FETP. Our ages range from 30s to 50s, and most of us identify as female. We are interested in learning processes and how to facilitate them with training approaches to improve public health and health security, and we are interested in how differences in cultural, educational, and professional backgrounds affect training and learning. Our positionality influences our perspective and relationships to the research and participants. Thus, we seek to co-design and co-interpret the study.

### Theoretical framework

We will situate this research in the constructivist paradigm with grounded theory, narrative inquiry, and critical action research. Grounded theory is a social research method that aims to generate explanatory theory inductively from analysis of social research data.(30) We will draw on the constructivist grounded theory methodology of Charmaz, who argues that while quantitative research aims for statistical inferences, grounded theory aims to fit theories emerging from the data with the data. It supplies evidence for emerging hypotheses that quantitative research could pursue.(31)

Narrative inquiry focuses on narratives, which display human existence situated in action.(32) It examines human lives and lived experiences as sources of legitimate knowledge and understanding (34) and reveals our social nature, social structures, and how humans make sense of the world(35). Here, we will employ Polkinghorne’s analysis of narratives and narrative analysis.(32)

As public health practitioners, we believe research should inform action. We will draw on the seminal work of Carr and Kemmis and of Wadsworth.(29, 33) Critical action research aims for change: explanations and understandings are not ends but steps in the process. Action research for learning relates practices, understandings, and situations to one another to discover correspondence and non-correspondence.(33)

### Setting and participants

Given the research resources available, we estimate that we can conduct 40 interviews and assume that fewer than eight interviews per program (2 trainers and 6 trainees of 3 per cohort) will not likely provide meaningful data. Thus, we aim to invite five programs to participate. Although including multiple geographical regions could provide useful insight, it will introduce more variability that will complicate interpretations. Working across geographical regions will also increase the cost of the research. Additionally, we consider that Basic and Frontline programs differ substantially from Intermediate and Advanced because of the training content, duration, and target participants (see 36, 37), and their inclusion will complicate interpretation. Thus, expecting that some invited programs will not participate, the PI has invited seven Intermediate and Advanced programs from the World Health Organization’s Western Pacific Region. These seven programs differ in duration, cohort size, years in operation, host institution, recruitment strategy, entry requirements, accreditation, and program design, as well as national language and health systems. They have similar aims for developing field epidemiologists through field-based service to contribute to national health security.

To participate, program directors will have to agree for trainers and trainees to enrol in the study and for one trainer to serve as a co-researcher committing to monthly virtual calls to discuss research aims, questions, direction, and interpretation of data. Among the invited programs, six have agreed to take part. At the time of writing, four will participate: Australia’s Master of Applied Epidemiology, Japan’s Field Epidemiology Training Program, Mongolia’s Field Epidemiology Training Program, and Taiwan’s Field Epidemiology Training Program. The other two programs expressed interest in joining the study but withdrew because of administrative issues.

Participants will include trainees and trainers in the participating programs. For this research, a ‘trainee’ will be anyone enrolled in a participating program during the study period. A ‘trainer’ for this research will be anyone recognized by the program as one who designs or delivers an activity intended to change the knowledge, skills, awareness, attitudes, or behaviours of trainees. In FETP, many people are involved in such activities: some are employed staff of the program with a responsibility to supervise or train; some are public health professionals not employed by the program who provide short courses, advise on projects, support field work, etc; and others lie between these extremes, such as professionals not employed by a program who supervise field deployments and lecture regularly. We will ask co-researchers to identify those most involved in training, mentoring, or supervising current trainees as ‘core trainers’. We will not exclude participants based on language.

We will conduct participant observations, during training and learning events in participating programs. Training and learning events will be those activities that are organized to intentionally change knowledge, skills, awareness, attitudes, or behaviours of field epidemiology trainees. We will utilize those events organized as a routine part of the program instead of requesting programs to organize events for observation. Examples include courses, workshops, case studies, mentor and supervisor meetings, and group projects. Because of the nature of participant observation, all trainers and trainees present during the observation will be participants, including trainers who may not be defined as ‘core trainers’, such as guest lecturers.

### Recruitment

Our sampling strategy will follow grounded theory approaches outlined by Glaser and Strauss and Charmaz.(31, 38) Glaser and Strauss promote ‘theoretical sampling’, a procedure with concurrent collection, coding, and analysis of data so that analyses can suggest further data to collect. They note that the researcher should select individuals and groups based on their potential to generate properties of categories and help relate categories to one another. For Charmaz, theoretical sampling means collecting more data about specific actions, experiences, events, and issues to illuminate variation within a category or process. Charmaz distinguishes initial sampling from theoretical sampling. For initial sampling, Charmaz advises setting criteria and starting with relevant material for the study. Theoretical sampling begins when preliminary categories have arisen and helps to check, qualify, and elaborate the categories and the relations among them.(31)

Our initial sampling will focus on participant observations and in-depth interviews. For participant observations, we will select one training and learning event per program in collaboration with each program’s co-researcher and program director. As the goal of the participant observation is to provide an initial view into the nature of the training-learning environment, the criterion for selecting the event is that it is routine for that program. To maximise resources, we will aim for events occurring over at least five consecutive days. The program director and co-researcher will distribute participant information sheets that are approved by the ethics review committee(s) and explain the participant observation to all individuals who may be present during the event, discuss risks and benefits, answer questions, and identify individuals who hesitate to participate. For these individuals, the director and co-researcher will ascertain if the hesitancy represents a refusal for the event to be observed or a refusal for data to be collected on that individual. In the case of the former, the participant observation will not proceed. For the later, the participant observation will proceed with no data being collected on that individual. Directors will then sign an assent form for the observation to proceed, which covers all participants at the event. All participants at the event will be advised that they may raise ethical concerns, hesitations, questions, and requests to halt the observation at any time by approaching the co-researcher, director, or PI.

For interviews, we will enrol a pool of trainers and trainees from which we can conduct initial and theoretical sampling. For trainees, we will use diverse strategies because the cohort size and cultures of participation in research varies across programs. For larger programs (>15 trainees) with high expected participation rates (>50% as judged by the co-researchers and PI), we will randomly select ten trainees to approach. For larger programs with low expected participation rates (<50%), as well as smaller programs (≤15 trainees) regardless of participation rates, we will approach all trainees. We will seek to enrol ‘core trainers’, and as the number of them is smaller than ten in each program we will approach all of them.

Co-researchers from the respective programs will send an email invitation to all selected individuals using the national language and include as attachments the participant information sheet and informed consent form approved by the ethics review committee(s). The invitation and information sheet will clarify that participation is voluntary and that no one from the respective programs knows if they have chosen to participate. The invitation will include a link to an enrolment website where the individual can enrol or decline to participate. For those choosing to enrol, the website will ask the participant to confirm having read the informed consent document and confirm agreement to participate with an electronic signature. When participants confirm agreement to participate, the website will request demographic information to facilitate initial sampling: year of birth, gender, program, language, role (trainer or trainee) and for trainers their number of years training. The registered responses will be available to the PI, not to the co-researchers. Because co-researchers will not know the responses of the invited individuals, they will send two follow-up emails at one-week intervals to all invited individuals in their programs. The follow-up email will say that the co-researcher does not know who has decided to enrol and thus is reminding all who have been invited. The pool of potential participants will comprise all those who indicate through the website that they agree to participate.

For initial sampling, we will aim to enrol from the pool three trainers and three trainees per cohort (e.g., six for Advanced programs) for each program. We believe three is a minimum initial sample to guide theoretical sampling. If two participants have opposing perspectives, a third perspective would suggest sampling towards one or the other. We will also aim to include one male, one female, one 35 or older, and one younger than 35 for both trainers and trainees from each program. Trained bi-lingual interviewers not associated with the programs will contact these individuals by email to arrange the first interviews. Interviews will begin with a review of informed consent, confirmation to video or audio record, and clarification that the participant can skip any question, halt the interview, and decide at any point until the data are prepared for publication to remove the interview from the study.

As we analyse data from the initial sample, categories and properties emerging from the data will suggest further sampling from the same participants (i.e., follow-up interviews) or from others who agreed to participate until we reach theoretical saturation. Charmaz describes theoretical saturation as the saturation of theoretical categories with data, when relationships among categories and variation within and between them have been defined, checked, and explained.(31) If necessary, we will invite trainers or trainees beyond the initial pool.

## Data collection

Data collection will employ two methods: participant observation and in-depth interviews. DeWalt and DeWalt describe participant observation as a data collection method that occurs in a naturalistic setting involving observation and/or participation in the activities of the people under study. As a method, it utilizes explicit recording and analysing of information gathered from participating and from observing. This method can provide context for sampling, interviewing, and construction of interview guides and is rarely the only technique for a study.(39) Patton summarises the advantages of the method: facilitates better understanding and capturing of context within which interactions occur; allows the researcher to be open, discovery-oriented, and inductive; and provides opportunities to see what escapes the awareness of the people in the setting and to learn about things that individuals may not disclose in interviews.(35)

For participant observations, the co-researcher of the respective program will introduce the PI at the beginning of the selected event, clarify procedures, and answer questions. The co-researcher will note that as an experienced field epidemiologist and trainer, the PI is available to support the event as much as possible. For example, trainers and trainees may ask the PI questions and solicit help where relevant. This approach aligns with the participant aspect of participant observation and reduces the discomfort of being observed by an outsider. The PI will participate in and observe participants’ interactions, discussions, and behaviours, as well as the setting. The PI will engage in casual conversations with participants letting them lead conversations to avoid tending towards interviews. Where the PI does not speak the language, a bi-lingual member of the program will interpret. The PI will record notes and convert them into field notes at the conclusion of each day. Words and behaviours will not be attributed to individual participants but to ‘a trainee’ or ‘a trainer.’ Field notes with the researcher’s ruminations on them will comprise the data of participant observations.

We will use in-depth interviews because, as Patton observes, they allow researchers to enter the perspective of research participants, understand what we have observed, and gather feelings, thoughts, intentions, and meaning.(35) Though we do not believe the topic is sensitive, participants in many FETP are government employees. They may hesitate to disclose opinions in participant observations or focus groups but feel comfortable doing so in confidential interviews.

We will conduct in-depth, unstructured interviews with trainees and trainers to explore experiences with training and learning in FETP. Interviews will last one to two hours and be conducted online. Participants can choose the time and place for the interviews according to their comfort and the rules of their workplace. For trainees, we will explore their experiences with training and learning. Training experiences have been defined above. Learning experiences will be those in which the trainee sought or perceived a change in knowledge, skills, attitudes, awareness, or behaviours for field epidemiology. Topics on the trainee initial interview guide include motivation to learn field epidemiology; daily life/world in the program; learning or conducting outbreak investigations, surveillance, public health communication, and epidemiologic methods; participating in courses or classes; mentoring and supervising; most difficult/important thing to learn in FETP; process for getting through struggles or challenges; perceived changes in self throughout FETP; how gender, age, culture, or background made the experience easier/more difficult; and how FETP fits with the trainee’s vision. For trainers, we will explore their experiences with training in these programs using similar topics on the trainer initial interview guide but relevant to training, e.g., motivation to train field epidemiologist.

We will train bi-lingual qualitative interviewers to conduct interviews in English, Japanese, Mongolian, or Mandarin Chinese, transcribe them, and translate them into English. Following the work of Polkinghorne(40) and McCance (41), we will train interviewers to use the guide to elicit trainees’ narratives of their experiences, emphasizing depth over breadth; to develop rapport with casual conversation; and to use broad questions based on the topics. For example, tell me about the last outbreak investigation you participated in. We will train them to avoid interrupting responses and to probe to facilitate relating stories.

Interviewers will transcribe the interviews. Interviewees will then have 30 days to review the transcripts to add comments, indicate where the meaning has been misunderstood, and request removal or alteration of sections that they feel are difficult to de-identify. This step will ensure participants do not perceive unnecessary risk to their program nor their career. Interviewers will then translate the interviews and include perceived changes in verbal and non-verbal communication, such as laughter and speeding up speech, to aid analyses.

We will digitise all data and upload them to secure password-protected folders. Once the PI has reviewed interview transcripts and the interviewer has clarified doubts and questions, we will destroy the original recordings.

### Data analysis

We will analyse the interview and participant observation data in three phases. For phase one, the PI will begin with open coding of data using the line-by-line comparison method (31) with NVivo 12 Pro (42). The PI will present the coding book and de-identified excerpts of the data to the co-researchers, who will then discuss interpretations as compared with those of the PI to focus analyses and inform phase two: focused coding. In focused coding, we will use constant comparison for larger sections of data and introduce analysis of narratives (32). Following Charmaz, we will retain multiple major codes while remaining open to modifying focused codes and moving between open and focused coding because the process is not linear.(31) We will employ Polkinghorne’s analysis of narratives, identifying stories about focused codes and using paradigmatic analysis to generate themes or classifications [categories and properties in grounded theory] across the stories.(32) The unit of analysis is thus the story in phase two, which Polkinghorne defines sustained and emplotted accounts having a beginning, middle, and end, plots being the conceptual schemes that display contextual meaning and draw events and actions together into an organized whole. During the third phase, we will employ Polkinghorne’s narrative analysis.(32) We will use the stories in the data and elements from data not in storied form to develop narratives that reflect the domains of interest to the researchers. We will integrate these narratives to reveal correspondence/non-correspondence of practices, understandings, and situations(33) for training and learning in FETP. We will arrange to dialogue with participating programs’ trainers and trainees including those not involved in interviews and with the wider FETP community to interpret correspondence and non-correspondence and explain training and learning.

### Trustworthiness and Credibility

We will implement the following procedures suggested by Patton to improve trustworthiness and credibility: 1) conduct in-depth qualitative field work guided by established methods and theoretical frameworks, integrating and triangulating diverse sources of qualitative data; 2) use constant comparative analyses; 3) involve multiple investigators to triangulate analyses; and 4) seek alternative explanations, divergent patterns, rival explanations, and negative cases in the data to avoid biases shaping the findings.(35) Also, following Wadsworth, the co-design, co-implementation, and co-interpretation of the research with practitioners in FETP should improve the study’s relevance, focus research questions, increase design effectiveness, improve meaningfulness of the data, strengthen power and accuracy of the generated theory, and improve the relevance and effectiveness of recommendations.(29)

## ETHICS AND DISSEMINATION

We have obtained approval from The Australian National University Human Research Ethics Committee to conduct this research (2021/771), unconditional approval from ANU Institutional Research, which oversees research on ANU students, and approval from the Institutional Review Board of Centers for Disease Control, Ministry of Health and Welfare, Taiwan (112206). No additional approvals are required.

We will publish findings in professional journals and conferences. A website with links to disseminated findings will be available to the public and to participants.

### Patient and Public Involvement Statement

As described in this, we have involved end-users as collaborative researchers in the design, implementation, and interpretation of the research and will engage program directors, trainers, and trainees including research participants to share and interpret findings.

## Data availability Statement

Data will not be publicly available because of privacy concerns.

## Data Availability

Data will not be publicly available because of privacy concerns.

## AUTHORS’ CONTRIBUTIONS

MMG developed the study concept and approach collaboratively with all co-authors, drafted the manuscript, and revised the manuscript.

EF developed the study concept and approach collaboratively with all co-authors and revised the manuscript.

ASH developed the study concept and approach collaboratively with all co-authors and reviewed the manuscript.

TS developed the study concept and approach collaboratively with all co-authors and revised the manuscript.

MB developed the study concept and approach collaboratively with all co-authors and reviewed the manuscript.

TH developed the study concept and approach collaboratively with all co-authors and revised the manuscript.

BP developed the study concept and approach collaboratively with all co-authors and revised the manuscript.

MDK developed the study concept and approach collaboratively with all co-authors and revised the manuscript.

## FUNDING STATEMENT

This research received no specific grant from any funding agency in the public, commercial or not-for-profit sectors.

## COMPETING INTERESTS STATEMENT

The authors declare no competing interests.

## Notes

### Competing Interest Statement

The authors have declared no competing interest.

## FULL REFERENCES

1. Parry AE, Law C, Pourmarzi D, Vogt F, Field E, Colquhoun S. Contribution of the Australian field epidemiology training workforce to the COVID-19 response, 2020. Western Pacific surveillance and response journal. 2022;13(4):1–100.

2. Singh SK, Dikid T, Dhuria M, Bahl A, Chandra R, Vaisakh TP, et al. India Field Epidemiology Training Program Response to COVID-19 Pandemic, 2020-2021. Emerging infectious diseases. 2022;28(13):S138–S44.

3. Cui A, Hamdani S, Woldetsadik MA, Clerville JW, Hu A, Abedi AA, et al. The Field Epidemiology Training Program’s Contribution to Essential Public Health Functions in Seven National Public Health Institutes. Int J Public Health. 2023;68:1606191.

4. Hu AE, Fontaine R, Turcios-Ruiz R, Abedi AA, Williams S, Hilmers A, et al. Field epidemiology training programs contribute to COVID-19 preparedness and response globally. BMC Public Health. 2022;22(1):63.

5. Griffith MM, Parry AE, Housen T, Stewart T, Kirk MD. COVID-19 and investment in applied epidemiology. Bull World Health Organ. 2022;100(7):415–a.

6. TEPHINET. About FETPs: TEPHINET; 2023 [Available from: https://www.tephinet.org/about/about-fetps.

7. U.S. Centers for Disease Control and Prevention. Field Epidemiology Training Program (FETP) 2021 [updated 17 December 2021. Available from: https://www.cdc.gov/globalhealth/healthprotection/fetp/index.htm.

8. Schaffner W, LaForce FM. Training field epidemiologists: Alexander D. Langmuir and the epidemic intelligence service. Am J Epidemiol. 1996;144(8 Suppl):S16–22.

9. Langmuir AD. The Epidemic Intelligence Service of the Center for Disease Control. Public Health Rep. 1980;95(5):470–7.

10. Etheridge EW. Sentinel for Health: A history of the Centers for Disease Control. Berkeley, CA: University of California Press; 1992.

11. Foege WH. Alexander D. Langmuir--his impact on public health. Am J Epidemiol. 1996;144(8 Suppl):S11–5.

12. Malison MD, Dayrit MM, Limpakarnjanarat K. The field epidemiology training programmes. International Journal of Epidemiology. 1989;18(4):995–6.

13. TEPHINET. Training Programs 2023 [updated May 2022. Available from: https://www.tephinet.org/training-programs.

14. Flint J. Evaluating the Impact of Field Epidemiology Training Programmes 2022 [Available from: https://static1.squarespace.com/static/5fb4723e225bcb20d28f0f76/t/609669cef4ebbb6e85bb50c0/1620470225427/Evaluating+the+Impact++of+Field+Epidemiology+Training+Programs.pdf.

15. Kirkpatrick DL, Kirkpatrick JD. Evaluating Training Programs: The Four Levels 3rd ed. San Francisco, CA: Berrett-Koehler Publishers, Inc. ; 2006.

16. Abduljalil M, Al Kohlani A, Jumaan A, Al Serouri A. Yemen Advanced Field Epidemiology Training Program: An Impact Evaluation, 2021. Epidemiologia (Basel). 2023;4(3):235–46.

17. Dey P, Brown J, Sandars J, Young Y, Ruggles R, Bracebridge S. The United Kingdom Field Epidemiology Training Programme: meeting programme objectives. Eurosurveillance. 2019;24(36).

18. Al Nsour M, Khader Y, Bashier H, Alsoukhni M. Evaluation of Advanced Field Epidemiology Training Programs in the Eastern Mediterranean Region: A Multi-Country Study. Frontiers in Public Health. 2021;9.

19. Cho KK, Marjadi B, Langendyk V, Hu W. The self-regulated learning of medical students in the clinical environment - a scoping review. BMC Med Educ. 2017;17(1):112.

20. Wiese A, Bennett D. Orientation of medical trainees to a new clinical environment (the ready-steady-go model): a constructivist grounded theory study. BMC Med Educ. 2022;22(1):37.

21. Kozato A, Shikino K, Matsuyama Y, Hayashi M, Kondo S, Uchida S, et al. A qualitative study examining the critical differences in the experience of and response to formative feedback by undergraduate medical students in Japan and the UK. BMC Med Educ. 2023;23(1):408.

22. Najafi Kalyani M, Jamshidi N, Molazem Z, Torabizadeh C, Sharif F. How do nursing students experience the clinical learning environment and respond to their experiences? A qualitative study. BMJ Open. 2019;9(7):e028052.

23. Lee JJ, Yang SC. Professional socialisation of nursing students in a collectivist culture: a qualitative study. BMC Med Educ. 2019;19(1):254.

24. Jantzen D. Refining nursing practice through workplace learning: A grounded theory. J Clin Nurs. 2019;28(13-14):2565–76.

25. Ohta R, Ryu Y, Katsube T, Otani J, Moriwaki Y. Strengths and Challenges for Medical Students and Residents in Rural Japan. Fam Med. 2021;53(1):32–8.

26. Collins A, Brown JS, Holum A. Cognitive apprenticeship: Making thinking visible. American educator. 1991;15(3):6–11.

27. Lave J, Wenger E. Situated learning : legitimate peripheral participation. Cambridge England ; New York: Cambridge University Press; 1991. 138 p. p.

28. Taber N, Plumb D, Jolemore S. “Grey” areas and “organized chaos” in emergency response. Journal of Workplace Learning. 2008;20(4):272–85.

29. Wadsworth Y. What is Participatory Action Research? Action research international. 1998:Paper 2.

30. Glaser BG, Strauss AL. The discovery of grounded theory; strategies for qualitative research. Chicago, IL: Aldine Publishing Co.; 1967.

31. Charmaz K. Constructing grounded theory. 2nd Edition. ed. London: SAGE; 2014.

32. Polkinghorne DE. Narrative configuration in qualitative analysis. International journal of qualitative studies in education. 1995;8(1):5–23.

33. Carr W, Kemmis S. Becoming Critical: Education, Knowledge and Action Research. London: The Falmer Press; 1986.

34. Clandinin DJ. Engaging in narrative inquiry. Walnut Creek, California: Left Coast Press, Inc; 2013.

35. Patton MQ. Qualitative research & evaluation methods : integrating theory and practice. Fourth edition. ed. Thousand Oaks, California: SAGE Publications, Inc.; 2015. xxi, 806 pages p. 36.

36. Andre AM, Lopez A, Perkins S, Lambert S, Chace L, Noudeke N, et al. Frontline Field Epidemiology Training Programs as a Strategy to Improve Disease Surveillance and Response. Emerg Infect Dis. 2017;23(13).

37. Traicoff DA, Suarez-Rangel G, Espinosa-Wilkins Y, Lopez A, Diaz A, Caceres V. Strong and Flexible: Developing a Three-Tiered Curriculum for the Regional Central America Field Epidemiology Training Program. Pedagogy in Health Promotion. 2015;1(2):74–82.

38. Glaser BG, Strauss AL. The discovery of grounded theory; strategies for qualitative research. Chicago,: Aldine Pub. Co.; 1967. x, 271 p. p.

39. DeWalt KM, DeWalt BR. Participant Observation : A Guide for Fieldworkers. Blue Ridge Summit, UNITED STATES: AltaMira Press; 2010.

40. Polkinghorne D. Narrative knowing and the human sciences. Albany: State University of New York Press; 1988.

41. McCance TV, McKenna HP, Boore JRP. Exploring caring using narrative methodology: an analysis of the approach. Journal of advanced nursing. 2001;33(3):350–6.

42. Lumivero. NVivo 12 Pro Windows. 12.6.1 ed 2017.

